# Development of an electronic frailty index for hospitalized geriatric patients in Sweden

**DOI:** 10.1101/2021.11.26.21266912

**Authors:** Jonathan K. L. Mak, Sara Hägg, Maria Eriksdotter, Martin Annetorp, Ralf Kuja-Halkola, Laura Kananen, Anne-Marie Boström, Miia Kivipelto, Carina Metzner, Viktoria Bäck Jerlardtz, Malin Engström, Peter Johnson, Lars Göran Lundberg, Elisabet Åkesson, Carina Sühl Öberg, Maria Olsson, Tommy Cederholm, Juulia Jylhävä, Dorota Religa

## Abstract

**Background:** Frailty assessment in the Swedish health system relies on the Clinical Frailty Scale (CFS), but it requires training, in-person evaluation, and is often missing in medical records. We aimed to develop an electronic frailty index (eFI) from routinely collected electronic health records (EHRs) and assess its predictive ability for adverse outcomes in geriatric patients.

**Methods:** EHRs were extracted for 18,225 geriatric patients with unplanned admissions between 1/3/2020 and 17/6/2021 from nine geriatric clinics in Stockholm, Sweden. A 48-item eFI was constructed using diagnostic codes, functioning and other health indicators, and laboratory data. The CFS, Hospital Frailty Risk Score, and Charlson Comorbidity Index were used for comparative assessment of the eFI. We modelled in-hospital mortality and 30-day readmission using logistic regression; 30-day and 6-month mortality using Cox regression; and length of stay using linear regression.

**Results:** 13,188 patients were included in analyses (mean age 83.1 years). A 10% increment in the eFI was associated with higher risks of in-hospital (odds ratio: 5.34; 95% confidence interval: 4.20-6.82), 30-day (hazard ratio [HR]: 3.28; 2.91-3.69), and 6-month mortality (HR: 2.70; 2.52-2.90) adjusted for age and sex. Of the frailty and comorbidity measures, the eFI had the best predictive accuracy for in-hospital mortality, yielding an area under receiver operating characteristic curve of 0.813. Higher eFI also predicted a longer length of stay, but had a rather poor discrimination for 30-day readmission.

**Conclusions:** An EHR-based eFI has good predictive accuracy for adverse outcomes, suggesting that it can be used in risk stratification in geriatric patients.

## INTRODUCTION

With the ageing of the global population, it becomes increasingly important to identify and support older adults who are at the greatest risk of adverse outcomes. Frailty, characterized by reduced physiological reserve and increased vulnerability to stressors (1), is a measure that captures such risk. Frailty has consistently been linked to negative health outcomes, such as mortality (2,3), hospitalization (4), and increased healthcare costs (5,6). Despite the lack of a universal consensus on how to best assess frailty, the most widely used and validated frailty models are the physical frailty phenotype (characterized by weight loss, weakness, slowness, inactivity, and exhaustion) (7) and the frailty index (FI; multidimensional deficit accumulation) (8). Nevertheless, these measures are time- and resource-consuming, which limits their incorporation into routine clinical practice (9). One of the most frequently adopted frailty measure in clinical settings is the Clinical Frailty Scale (CFS) (10), which is a quick and simple screening tool and often has high accuracy and feasibility (11). However, as an instrument that requires in-person assessment, it may be subject to interrater error and may not always be available at hospital admission (12–14).

Automated frailty scores, based on routinely collected electronic health records (EHRs) or administrative claims data, have recently been developed in certain countries such as the UK (15), the US (16,17), and Canada (18). One of the first models was the electronic frailty index (eFI) proposed by Clegg *et al*. (15), who created it using the UK primary care Read codes on the basis of the Rockwood deficit accumulation approach (8). Several variations of the eFI, such as claims-based frailty indices (16,18), or an eFI incorporating diagnosis codes, functional impairments, and laboratory measures (17), have later been developed. While these tools are commonly used on data retrieved from primary care, recent studies have shown that such eFIs show good predictive performance for all-cause mortality also in hospital settings (19–21). Another database-derived frailty measure, the Hospital Frailty Risk Score (HFRS), is calculated based on the *International Classification of Diseases, Tenth Revision* (ICD-10) codes (22), and has been validated for its ability to predict mortality and prolonged length of stay in hospitalized older patients (23–25). However, its composition of ICD-10 diagnoses makes it more similar to a comorbidity measure, possibly missing out other frailty aspects such as functioning (22).

To date, frailty is not yet routinely assessed in all older adults in Sweden, but the CFS has started to be implemented in Stockholm, at least in geriatric clinics. Routine frailty screening can help identifying patients who would benefit most from a Comprehensive Geriatric Assessment (CGA) (26). To reduce the burden of bedside frailty assessment and aid in risk stratification, there is also an increasing need to analyze whether an eFI can be adapted to Swedish context. The objective of this study was therefore to develop an eFI using routinely collected EHRs in geriatric clinics in Stockholm, Sweden. For validation, we compared its predictive ability to other validated frailty and comorbidity measures, the CFS, HFRS, and the Charlson Comorbidity Index (CCI) (27), considering mortality, readmission and the length of stay as outcomes.

## METHODS

### Study design and sample

We conducted a retrospective cohort study using electronic medical records in the Stockholm Region. Between 1 March 2020 and 17 June 2021, patients with unplanned admissions to nine geriatric clinics treated for any causes, except COVID-19, were included. COVID-19 patients (defined based on ICD-10 codes of U07.1, U07.2, U08.9, U09.9, or U10.9) often have different characteristics and prognosis and were analyzed in a separate paper. Exclusion criteria included admissions without discharge information or with a length of stay <24h. For patients with multiple admissions during the study period, the first available admission was considered in the analyses. In total, 18,225 non-COVID-19 geriatric patients were included, of whom 13,188 (72.4%) had sufficient data for calculation of the eFI. A flowchart of sample selection is presented in **Supplementary Figure 1**. This study was approved by the Swedish Ethical Review Authority (Dnr 2020-02146, 2020-03345, 2021- 00595, 2021-02096).

### Construction of the electronic frailty index

Following the eFI model developed by Pajewski *et al*. (17), we selected a total of 48 items for the construction of the eFI. The items consisted of deficits in three categories: (i) disease diagnoses, extracted from ICD-10 codes assigned during the current admission (29 items), (ii) functional data and other health indicators, such as falls, neuropsychological problems, and oral health (10 items), and (iii) laboratory measures (9 items). The full list of the eFI items and the coding methods are shown in **Supplementary Table 1**. Consistent with the original deficit accumulation model (8), we calculated the eFI as the sum of deficit items divided by the total number, providing that the patient had ≥30 non-missing items (8). For instance, a patient with 10 deficit points out of 45 non-missing items would receive an eFI of 10/45=0.22. As 60% (29 out of 48) of the eFI items were based on ICD-10 codes (which were non-missing for all participants), we required that at least half of the functioning and/or laboratory-based deficit items were non-missing in each individual. This was to avoid the lack of such items that are necessary in reaching the multidimensional definition of frailty, and prevent the eFI from having an overrepresentation of comorbidity items. Based on the distribution of the eFI in our sample and by adopting a previously used cut-off of 0.25 (28,29), we categorized patients into four groups: fit (eFI ≤0.15), mild frailty (0.15> eFI ≥0.2), moderate frailty (0.2> eFI ≥0.25), and severe frailty (eFI >0.25).

### Other frailty and comorbidity measures

Other frailty measures, the CFS and HFRS, and the CCI, which is a measure of comorbidity, were used for comparative performance assessment and validation of the eFI. The CFS was scored by a physician or a trained nurse during first day of admission, using a chart review and face-to-face assessments. The CFS ranges from 1 (“very fit”) to 9 (“terminally ill”), and was categorized in three groups: CFS 1–3, CFS 4–5, CFS 6–9. Both the HFRS and CCI were calculated based on ICD-10 codes. The HFRS is based on 109 frailty-associated and differently weighted ICD-10 codes, and categorizes the individuals into low (<5), intermediate (5–15) and high risk (>15) frailty groups (22). The CCI was computed using an algorithm adapted for Swedish settings (30), and was considered as a continuous variable in all analyses.

### Outcomes

Primary outcomes were in-hospital, 30-day, and 6-month all-cause mortality, calculated from the date of admission. Dates of death were extracted from the Population System in Sweden. Secondary outcomes were 30-day readmission to any of the nine included clinics, and the length of stay. Only patients discharged to home were included in the 30-day readmission analysis, i.e., excluding those who were transferred to another unit or care facility after the stay at the geriatric unit.

### Statistical analysis

Patients’ characteristics were summarized and compared by sex using *t*-tests or Mann-Whitney *U* tests for continuous variables, and Pearson *χ*^2^ tests for categorical variables. Spearman’s correlations (*ρ*) were calculated between continuous frailty and comorbidity measures.

We fitted multivariable regression models to compare the eFI to the CFS, HFRS, and CCI in predicting different outcomes, adjusting for age and sex in all models. In-hospital mortality and 30-day readmission were modelled using logistic regression, with odds ratios (ORs) and 95% confidence intervals (CI) presented. The predictive accuracy (discrimination) of the logistic regression models was examined using area under the receiver operating characteristic curve (AUC). Hazard ratios (HRs) and 95% CI for 30-day and 6-month mortality were estimated using Cox proportional-hazards models. Harrell’s C-statistics were calculated for assessing predictive accuracy of Cox models, with 95% CI calculated through 1,000 bootstrapping resampling. Linear regression models were performed for the length of stay, and coefficients of determination (*R*^2^) were calculated to assess the proportion of variation explained by the independent variables.

As a sensitivity analysis, we stratified the analysis of in-hospital mortality by the nine admitting clinics to assess the variation by hospitals.

All analyses were performed using R version 4.0.5.

## RESULTS

### Sample characteristics

Among the 13,188 patients who had sufficient data for calculation of the eFI, the mean age was 83.1 years and 60.2% were women; the in-hospital mortality rate was 1.4% and the median length of stay was 6.7 days (**Table 1**). The patients with and without sufficient data for calculation of the eFI were similar with regards to age, sex, frailty level according to CFS and HFRS, and in-hospital death rate (**Supplementary Table 2**).

**Table 1.** Sample characteristics by sex among patients with the electronic frailty index available

| Characteristic | All patients<br>(n = 13,188) | Women<br>(n = 7,943) | Men<br>(n = 5,245) | *p*<sup>a</sup> |
| --- | --- | --- | --- | --- |
| Age, year, mean ± SD | 83.1 ± 8.4 | 84.1 ± 8.3 | 81.6 ± 8.4 | <0.001 |
| Age category, n (%) |  |  |  | <0.001 |
| <65 years | 215 (1.6) | 91 (1.1) | 124 (2.4) |  |
| 65–74 years | 1,917 (14.5) | 995 (12.5) | 922 (17.6) |  |
| 75–84 years | 4,909 (37.2) | 2,764 (34.8) | 2,145 (40.9) |  |
| 85–94 years | 5,103 (38.7) | 3,319 (41.8) | 1,784 (34.0) |  |
| ≥95 years | 1,044 (7.9) | 774 (9.7) | 270 (5.1) |  |
| eFI, median [IQR] | 0.181 [0.143, 0.222] | 0.179 [0.141, 0.220] | 0.185 [0.145, 0.227] | <0.001 |
| eFI category, n (%) |  |  |  | <0.001 |
| Fit (≤0.15) | 3,860 (29.3) | 2,422 (30.5) | 1,438 (27.4) |  |
| Mild frailty (>0.15–0.2) | 4,366 (33.1) | 2,650 (33.4) | 1,716 (32.7) |  |
| Moderate frailty (>0.2–0.25) | 3,220 (24.4) | 1,896 (23.9) | 1,324 (25.2) |  |
| Severe frailty (>0.25) | 1,742 (13.2) | 975 (12.3) | 767 (14.6) |  |
| CFS score, median [IQR] | 5 [4, 6] | 5 [4, 6] | 5 [4, 6] | 0.99 |
| CFS category, n (%) |  |  |  | 0.87 |
| 1–3 | 830 (6.3) | 508 (6.4) | 322 (6.1) |  |
| 4–5 | 1,869 (14.2) | 1,117 (14.1) | 752 (14.3) |  |
| 6–9 | 2,246 (17.0) | 1,362 (17.2) | 884 (16.9) |  |
| Missing | 8,243 (62.5) | 4,956 (62.4) | 3,287 (62.7) |  |
| HFRS, median [IQR] | 2.8 [1.4, 5.0] | 2.6 [1.3, 4.8] | 3.0 [1.4, 5.2] | <0.001 |
| HFRS category, n (%) |  |  |  | <0.001 |
| Low risk (<5) | 9,811 (74.4) | 6,033 (76.0) | 3,778 (72.0) |  |
| Intermediate risk (5–15) | 3,316 (25.1) | 1,881 (23.7) | 1,435 (27.4) |  |
| High risk (>15) | 61 (0.5) | 29 (0.4) | 32 (0.6) |  |
| CCI, median [IQR] | 1 [0, 2] | 1 [0, 2] | 1 [0, 3] | <0.001 |
| In-hospital mortality, n (%) | 183 (1.4) | 96 (1.2) | 87 (1.7) | 0.037 |
| Discharged to home, n (%) | 10,448 (79.2) | 6,330 (79.7) | 4,118 (78.5) | 0.11 |
| 30-day readmission<sup>b</sup>, n (%) | 1,114 (10.7) | 672 (10.6) | 442 (10.7) | 0.88 |
| Length of stay, day, median [IQR] | 6.7 [4.7, 8.9] | 6.7 [4.7, 8.9] | 6.5 [4.6, 8.8] | 0.022 |

Note: CCI, Charlson Comorbidity Index; CFS, Clinical Frailty Scale; eFI, electronic frailty index; HFRS, Hospital Frailty Risk Score; IQR, interquartile range; SD, standard deviation
<sup>a</sup> *P*-values for comparison between sexes, based on *t*-tests or Mann-Whitney *U* tests for continuous variables, and $\chi^2$ tests for categorical variables
<sup>b</sup> Only patients discharged to home after the first admission were included for analysis of 30-day readmission

The eFI was approximately normally distributed, with a median of 0.181 (interquartile range: 0.143–0.222; range: 0–0.432) (**Supplementary Figure 2**). The proportions of patients classified as fit, mildly frail, moderately frail, and severely frail were 29.3%, 33.1%, 24.4%, and 13.2%, respectively. Although women were on average older than men (mean age: 84.1 vs. 81.6), men had significantly higher frailty scores than women when defined by the eFI (severe frailty: 14.6% vs. 12.3%) and the HFRS (intermediate or high risk: 28.0% vs. 24.1%), but not by the CFS (**Table 1**). The eFI was moderately correlated with the CFS (*ρ*=0.420), and to a lesser extent with the HFRS (*ρ*=0.289) and CCI (*ρ*=0.368) (**Supplementary Figure 3 & Supplementary Table 3**).

### Associations with in-hospital, 30-day, and 6-month mortality

After adjusting for age and sex, the eFI was strongly associated with in-hospital mortality (OR per 10% increase: 5.34, 95% CI: 4.20–6.82) (**Table 2**). The CFS, HFRS, and CCI also had positive associations with in-hospital mortality; however, among all the measures, the eFI had the greatest predictive accuracy for in-hospital mortality when added to a model with age and sex, yielding an AUC of 0.813 (95% CI: 0.769–0.857) (**Figure 1A**). As a sensitivity analysis, we stratified the sample by the nine clinics and found robust predictive performance of the eFI for in-hospital mortality across the clinics, with AUCs ranging from 0.745–0.871 (**Supplementary Table 4**). The associations were also essentially unchanged when included the admitting clinics as a covariate in the models, or stratified by sex and age (data not shown).

**Table 2.**
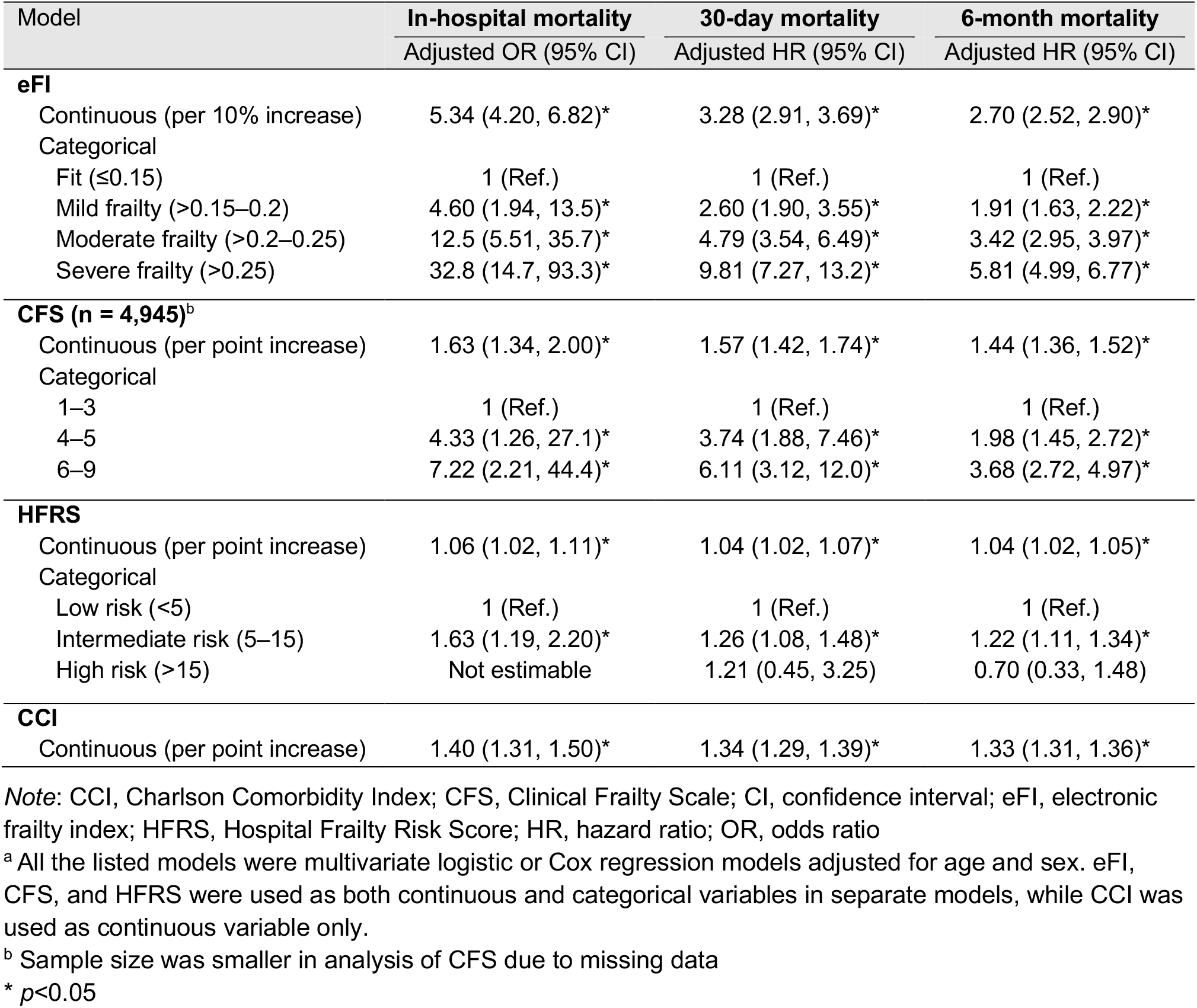
Associations between frailty and comorbidity measures and mortality outcomes (n = 13,188)^a^

| Model | In-hospital mortality<br>Adjusted OR (95% CI) | 30-day mortality<br>Adjusted HR (95% CI) | 6-month mortality<br>Adjusted HR (95% CI) |
| --- | --- | --- | --- |
| **eFI** |  |  |  |
| Continuous (per 10% increase) | 5.34 (4.20, 6.82)* | 3.28 (2.91, 3.69)* | 2.70 (2.52, 2.90)* |
| Categorical |  |  |  |
| Fit ($\leq 0.15$) | 1 (Ref.) | 1 (Ref.) | 1 (Ref.) |
| Mild frailty ($>0.15-0.2$) | 4.60 (1.94, 13.5)* | 2.60 (1.90, 3.55)* | 1.91 (1.63, 2.22)* |
| Moderate frailty ($>0.2-0.25$) | 12.5 (5.51, 35.7)* | 4.79 (3.54, 6.49)* | 3.42 (2.95, 3.97)* |
| Severe frailty ($>0.25$) | 32.8 (14.7, 93.3)* | 9.81 (7.27, 13.2)* | 5.81 (4.99, 6.77)* |
| **CFS (n = 4,945)<sup>b</sup>** |  |  |  |
| Continuous (per point increase) | 1.63 (1.34, 2.00)* | 1.57 (1.42, 1.74)* | 1.44 (1.36, 1.52)* |
| Categorical |  |  |  |
| 1–3 | 1 (Ref.) | 1 (Ref.) | 1 (Ref.) |
| 4–5 | 4.33 (1.26, 27.1)* | 3.74 (1.88, 7.46)* | 1.98 (1.45, 2.72)* |
| 6–9 | 7.22 (2.21, 44.4)* | 6.11 (3.12, 12.0)* | 3.68 (2.72, 4.97)* |
| **HFRS** |  |  |  |
| Continuous (per point increase) | 1.06 (1.02, 1.11)* | 1.04 (1.02, 1.07)* | 1.04 (1.02, 1.05)* |
| Categorical |  |  |  |
| Low risk ($<5$) | 1 (Ref.) | 1 (Ref.) | 1 (Ref.) |
| Intermediate risk (5–15) | 1.63 (1.19, 2.20)* | 1.26 (1.08, 1.48)* | 1.22 (1.11, 1.34)* |
| High risk ($>15$) | Not estimable | 1.21 (0.45, 3.25) | 0.70 (0.33, 1.48) |
| **CCI** |  |  |  |
| Continuous (per point increase) | 1.40 (1.31, 1.50)* | 1.34 (1.29, 1.39)* | 1.33 (1.31, 1.36)* |

Note: CCI, Charlson Comorbidity Index; CFS, Clinical Frailty Scale; CI, confidence interval; eFI, electronic frailty index; HFRS, Hospital Frailty Risk Score; HR, hazard ratio; OR, odds ratio
<sup>a</sup> All the listed models were multivariate logistic or Cox regression models adjusted for age and sex. eFI, CFS, and HFRS were used as both continuous and categorical variables in separate models, while CCI was used as continuous variable only.
<sup>b</sup> Sample size was smaller in analysis of CFS due to missing data
\* $p < 0.05$

**Figure 1.**
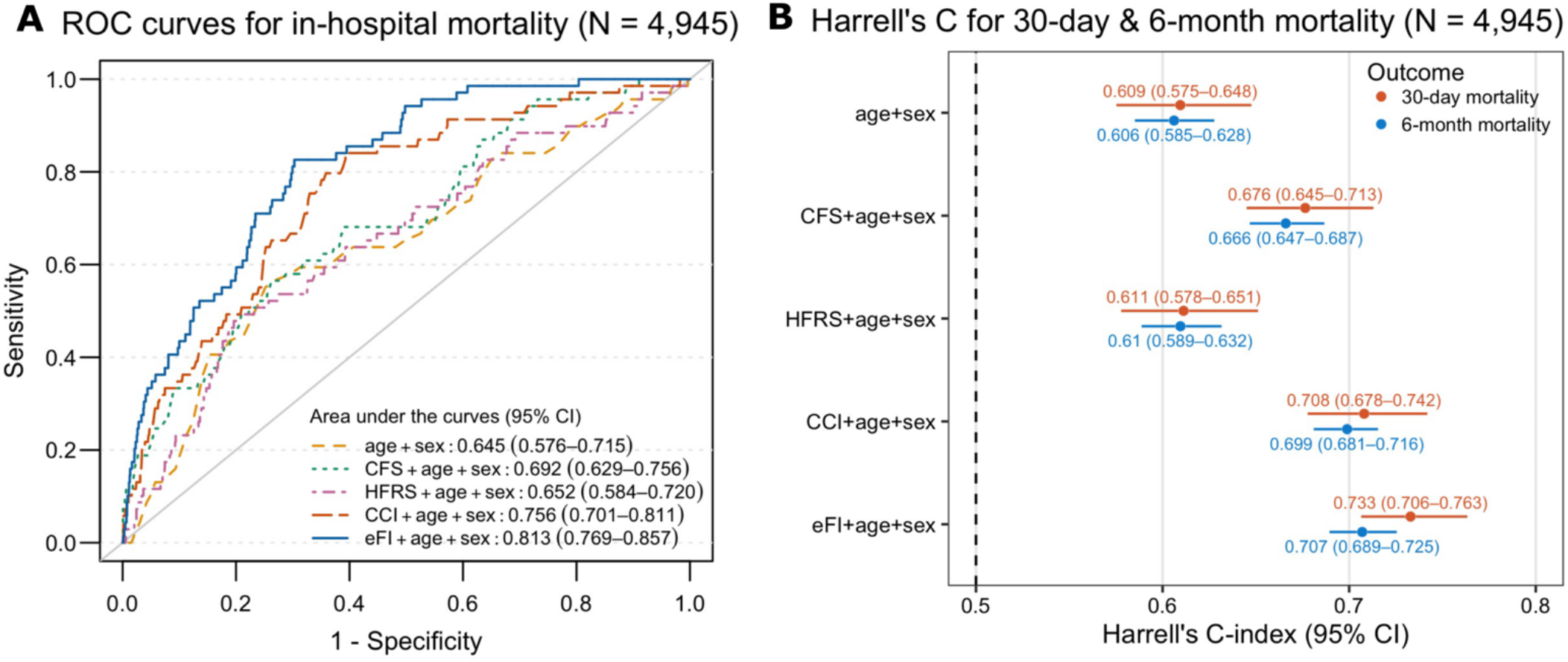
Predictive accuracies of frailty and comorbidity measures for mortality outcomes in patients with all measures available (n = 4,945). **(A)** Receiver operating characteristics (ROC) curves for in-hospital mortality; **(B)** Harrell’s C-statistics from Cox models for 30-day and 6-month mortality. CFS, HFRS, CCI, and eFI were considered as continuous variables in all the models. Abbreviations: CI, confidence interval; CFS, Clinical Frailty Scale; HFRS, Hospital Frailty Risk Score; CCI, Charlson Comorbidity Index; eFI, electronic frailty index

We observed a gradient of an increasing risk of mortality in the higher eFI categories compared to the lowest category (fit) in six months from admission (**Figure 2**). Similar to in-hospital mortality, a 10% rise in the eFI score was significantly associated with elevated risks of 30-day (HR: 3.28, 95% CI: 2.91–3.69) and 6-month mortality (HR: 2.70, 95% CI: 2.52– 2.90) after adjusting for age and sex (**Table 2**). The eFI models had the highest Harrell’s C-statistics (0.733 for 30-day mortality & 0.707 for 6-month mortality) in comparison to the other frailty and comorbidity measures, indicating that the eFI had good predictive accuracy for 30-day and 6-month mortality in the Cox models (**Figure 1B**).

**Figure 2.**
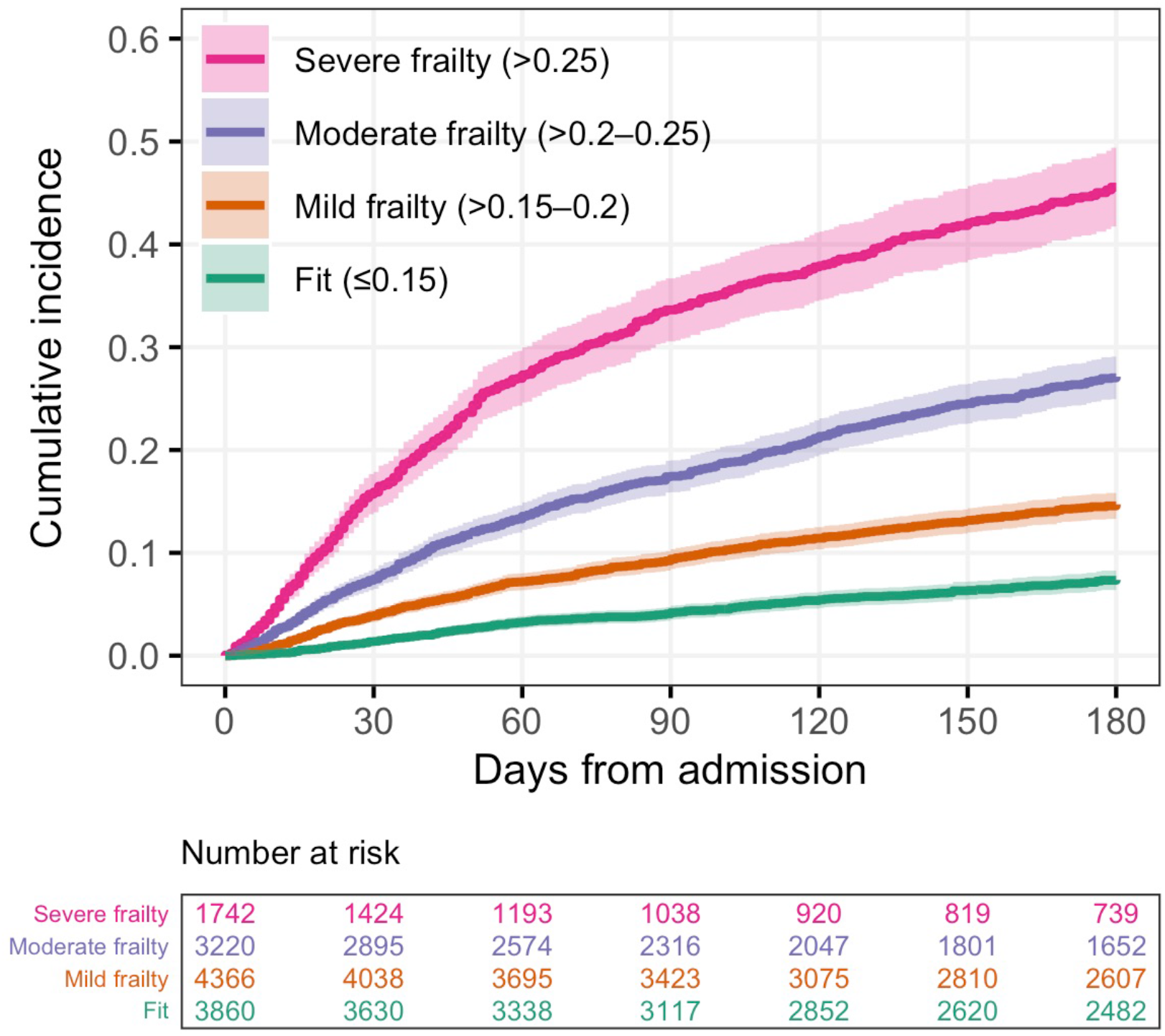
Kaplan-Meier curves for all-cause mortality by categories of the electronic frailty index (n = 13,188)

### Associations with 30-day readmission and length of stay

Despite the significant associations between the eFI and 30-day readmission (OR per 10% increase: 1.50, 95% CI: 1.35–1.68), and likewise between the CFS, HFRS and CCI, and 30-day readmission, the AUCs for these models were <0.6, indicating poor discrimination for readmission (**Supplementary Table 5**).

There was an approximately linear relationship between the eFI and the length of stay, regardless of age and sex (**Figure 3**). Similarly, linear regression analysis showed that after adjusting for age and sex, a 10% increment in the eFI was on average associated with a 2-days longer hospital stay (95% CI: 1.88–2.13). The model including eFI, age, and sex explained 7.21% of the variation in the length of stay (**Supplementary Table 6**).

**Figure 3.**
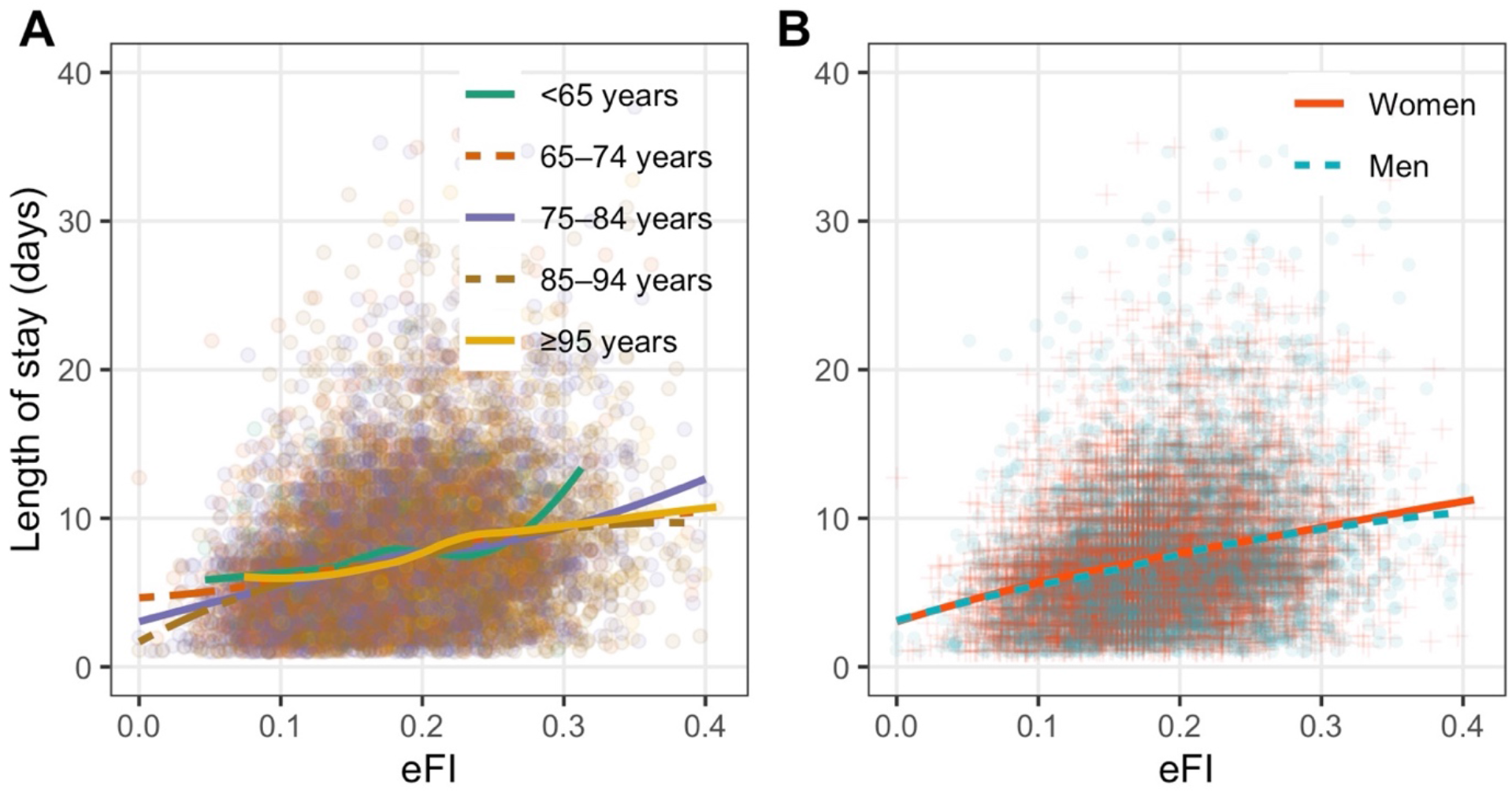
Scatter plots of the electronic frailty index and the length of stay (n = 13,188). **(A)** stratified by age; **(B)** stratified by sex. The colored fit lines represent the fitted locally estimated scatterplot smoothing curves (LOESS).

## DISCUSSION

Although it is well-known that frailty predicts adverse outcomes, traditional frailty measurements often require in-person assessment and may not always be feasible or available in clinical settings. There is a growing interest of deriving frailty measures from routinely collected health data to facilitate frailty screening in clinical practice. Several such eFI models have been developed for primary care, and inpatient and outpatient settings using diagnostic codes, health service codes, and clinical information (31). Here we adapted the US eFI model (17) to Swedish EHR data, i.e. retrieved from geriatric clinics in the Stockholm region. We found that the eFI outperformed the currently available frailty and comorbidity scales (i.e., CFS, HFRS, and CCI) in predicting in-hospital mortality, yielding an AUC of 0.813 when added to a model with age and sex. It also had better accuracy for 30-day and 6-month mortality than CFS and HFRS and was significantly associated with longer length of stay. Nevertheless, all the frailty scales and the CCI had a poor predictive accuracy for 30-day readmission.

Based on data availability and following the model developed by Pajewski *et al*. (17), we included ICD-10 codes, functional assessment scales and other health indicators, and laboratory measurements in the eFI. It had a moderate correlation with a clinical frailty measure (i.e., CFS, *ρ*=0.420), but a weaker correlation with a comorbidity measure (i.e., CCI, *ρ*=0.368) and a frailty measure based on ICD-10 codes (i.e., HFRS, *ρ*=0.289). These findings suggest that the eFI captures frailty rather than multimorbidity. This is also on par with previous studies showing a moderate correlation for the eFI with the CFS (32), but a low correlation with the HFRS (33).

Although a data-driven or machine learning approach can be used in claims-based frailty indices (31), we applied the conventional deficit accumulation model (8) – a generalizable approach to frailty, in which a wide range of deficits e.g., signs, symptoms, diseases, functional limitations, laboratory measures, can be included as long as it includes at least 30 age-related deficits (8,34). We noticed, however, some differences in the characteristics of our eFI compared to a survey-based Rockwood FI. First, instead of a right-skewed distribution characteristic to the Rockwood FI, our eFI was approximately normally distributed, possibly due to a homogenous sample of less healthy hospitalized older adults (35,36), and inclusion of laboratory tests (representing subclinical and cellular deficits) (37,38). Second, our eFI had a relatively low maximum value of 0.432. One possible reason is that for assessing the diagnosis-based ICD-code items (29 out of 48), we used only those codes that were recorded during the current visit, possibly leading to underreporting of certain diseases. Third, we observed slightly, but significantly higher eFI scores in men than in women. Although FI scores are generally higher in women in community populations (39), no sex difference (36,40) or higher scores in men (18) have been found in hospitalized patients. The sex differences may also be influenced by whether self-reported (41) and laboratory deficits (37) are included. Indeed, more studies are needed to understand the mechanisms regarding sex differences in frailty (39).

Our finding of a robust predictive ability of the eFI for mortality and the length of stay is consistent with previous studies (18–21). Importantly, our eFI performed better than the CFS in predicting adverse outcomes, hence providing important insights on its potential use in risk stratification in geriatric patients to ease personnel burden in hospital settings. However, all the frailty and comorbidity measures had a poor predictive accuracy for 30-day readmission. This may partly be explained by the potential misclassification of readmission, since we did not have data on patients who were admitted to other clinics than the nine geriatric clinics. Besides, a relatively low AUC or c-statistic for frailty in predicting readmission (around 0.5–0.6) is also frequently reported in the literature (19,22,23,33,42,43). Hospital readmission is often described as a complex outcome not merely related to the health status of patients, but also social factors such as access to care, social support, and drug abuse (44,45), which are factors not captured in the frailty measures. We may speculate that many of the more frail individuals resided in elderly care homes where care facilities are better, have a less pronounced tendency for readmission to hospital, compared to less frail individuals living in their own homes.

This study has a relatively large sample size from multiple hospitals, allowing us to assess the predictive performance of the eFI across the nine clinics. We also had data on other validated frailty and comorbidity measures for validation and comparative assessment of the eFI. Nevertheless, some limitations should be considered. As mentioned above, the use of ICD-10 codes from a single admission may have caused an underreporting of the diseases, and the same applies to the HFRS and CCI. As we only analyzed patients in geriatric clinics, our results cannot be applied to older patients in other units. It would be of interest to investigate whether the eFI performs equally well in other older in-patient groups. Finally, the eFI could not be calculated for 27.6% of our sample due to missing data on functioning and laboratory measures. Our rate of missing data is nevertheless comparable to the ∼30% missing data in the US eFI by Pajewski *et al*. (17). Instead of the patients’ demographic characteristics and health status being associated with the rate of missing data in the US eFI, our missing data were more related to variations in the data collection practice between hospitals. The COVID-19 pandemic might have affected the data collection routines for the functional and other health assessments, leading to more missing data than usual. Future work may reveal which eFI items are the most decisive for predicting adverse outcomes, as well as which alternative available items from the EHRs could be included to complement the current eFI. Furthermore, it may be relevant to use other outcome measures than mortality in coming research efforts, although measures related to physical function and cognition are inherent in the FI. Crucial is also to show whether the eFI is able to identify patients that will benefit from CGA (26), and that will respond to treatment tailored from the outcome of the CGA. When implementing these results, it is also essential to include patients’ perspectives and to understand their feelings on frailty (46,47).

To conclude, we developed an eFI based on routinely collected EHRs for Swedish geriatric patients, and showed that it correlates with the CFS and has a good predictive accuracy for short-and long-term mortality and the length of stay. This work provides evidence that an eFI is feasible for risk stratification among geriatric patients in Stockholm, Sweden, and may as well be applicable to countries with similar health system.

## Data Availability

All data produced in the present study are available upon reasonable request to the authors.

## ACKNOWLEDGEMENTS

We would like to thank all patients, caregivers and staff who contributed to this study. This study was accomplished within the context of the Swedish National Graduate School for Competitive Science on Ageing and Health (SWEAH) funded by the Swedish Research Council.

## REFERENCES

1. Hoogendijk EO, Afilalo J, Ensrud KE, Kowal P, Onder G, Fried LP. Frailty: implications for clinical practice and public health. Lancet. 2019;394(10206):1365–1375. doi:10.1016/S0140-6736(19)31786-6.

2. Kojima G, Iliffe S, Walters K. Frailty index as a predictor of mortality: A systematic review and meta-analysis. Age Ageing. 2018;47(2):193–200. doi:10.1093/ageing/afx162.

3. Chong E, Ho E, Baldevarona-Llego J, Chan M, Wu L, Tay L. Frailty and risk of adverse outcomes in hospitalized older adults: a comparison of different frailty measures. J Am Med Dir Assoc. 2017;18(7):638.e7–638.e11. doi:10.1016/j.jamda.2017.04.011.

4. Chang S-F, Lin H-C, Cheng C-L. The relationship of frailty and hospitalization among older people: evidence from a meta-analysis. J Nurs Scholarsh. 2018;50(4):383–391. doi:https://doi.org/10.1111/jnu.12397.

5. Hajek A, Bock J-O, Saum K-U, et al. Frailty and healthcare costs—longitudinal results of a prospective cohort study. Age Ageing. 2018;47(2):233–241. doi:10.1093/ageing/afx157.

6. García-Nogueras I, Aranda-Reneo I, Peña-Longobardo LM, Oliva-Moreno J, Abizanda P. Use of health resources and healthcare costs associated with frailty: The FRADEA study. J Nutr Health Aging. 2017;21(2):207–214. doi:10.1007/s12603-016-0727-9.

7. Fried LP, Tangen CM, Walston J, et al. Frailty in older adults: evidence for a phenotype. Journals Gerontol Ser A Biol Sci Med Sci. 2001;56(3):M146–M157. doi:10.1093/gerona/56.3.m146.

8. Searle SD, Mitnitski A, Gahbauer EA, Gill TM, Rockwood K. A standard procedure for creating a frailty index. BMC Geriatr. 2008;8(1):24. doi:10.1186/1471-2318-8-24.

9. Dent E, Kowal P, Hoogendijk EO. Frailty measurement in research and clinical practice: A review. Eur J Intern Med. 2016;31:3–10. doi:10.1016/j.ejim.2016.03.007.

10. Rockwood K, Song X, MacKnight C, et al. A global clinical measure of fitness and frailty in elderly people. CMAJ. 2005;173(5):489–495. doi:10.1503/cmaj.050051.

11. Aucoin SD, Hao M, Sohi R, et al. Accuracy and feasibility of clinically applied frailty instruments before surgery: a systematic review and meta-analysis. Anesthesiology. 2020;133(1):78–95. doi:10.1097/ALN.0000000000003257.

12. Shrier W, Dewar C, Parrella P, Hunt D, Hodgson LE. Agreement and predictive value of the Rockwood Clinical Frailty Scale at emergency department triage. Emerg Med J. November 2020:emermed-2019-208633. doi:10.1136/emermed-2019-208633.

13. Surkan M, Rajabali N, Bagshaw SM, Wang X, Rolfson D. Interrater reliability of the Clinical Frailty Scale by geriatrician and intensivist in patients admitted to the intensive care unit. Can Geriatr J. 2020;23(3):235–241. doi:10.5770/cgj.23.398.

14. Pugh RJ, Battle CE, Thorpe C, et al. Reliability of frailty assessment in the critically ill: a multicentre prospective observational study. Anaesthesia. 2019;74(6):758–764. doi:10.1111/anae.14596.

15. Clegg A, Bates C, Young J, et al. Development and validation of an electronic frailty index using routine primary care electronic health record data. Age Ageing. 2016;45(3):353–360. doi:10.1093/ageing/afw039.

16. Kim DH, Schneeweiss S, Glynn RJ, Lipsitz LA, Rockwood K, Avorn J. Measuring frailty in Medicare data: development and validation of a claims-based frailty index. Journals Gerontol Ser A. 2018;73(7):980–987. doi:10.1093/gerona/glx229.

17. Pajewski NM, Lenoir K, Wells BJ, Williamson JD, Callahan KE. Frailty screening using the electronic health record within a Medicare accountable care organization. Journals Gerontol Ser A. 2019;74(11):1771–1777. doi:10.1093/gerona/glz017.

18. McIsaac DI, Wong CA, Huang A, Moloo H, van Walraven C. Derivation and validation of a generalizable preoperative frailty index using population-based health administrative data. Ann Surg. 2019;270(1):102–108. doi:10.1097/SLA.0000000000002769.

19. Callahan KE, Clark CJ, Edwards AF, et al. Automated frailty screening at-scale for pre-operative risk stratification using the electronic frailty index. J Am Geriatr Soc. 2021;69(5):1357–1362. doi:10.1111/jgs.17027.

20. Tew YY, Chan JH, Keeling P, et al. Predicting readmission and death after hospital discharge: a comparison of conventional frailty measurement with an electronic health record-based score. Age Ageing. March 2021. doi:10.1093/ageing/afab043.

21. Darvall JN, Greentree K, Loth J, et al. Development of a frailty index from routine hospital data in perioperative and critical care. J Am Geriatr Soc. 2020;68(12):2831–2838. doi:https://doi.org/10.1111/jgs.16788.

22. Gilbert T, Neuburger J, Kraindler J, et al. Development and validation of a Hospital Frailty Risk Score focusing on older people in acute care settings using electronic hospital records: an observational study. Lancet. 2018;391(10132):1775–1782. doi:10.1016/S0140-6736(18)30668-8.

23. Gilbert T, Cordier Q, Polazzi S, et al. External validation of the Hospital Frailty Risk Score in France. Age Ageing. June 2021. doi:10.1093/ageing/afab126.

24. Harvey LA, Toson B, Norris C, Harris IA, Gandy RC, Close JJCT. Does identifying frailty from ICD-10 coded data on hospital admission improve prediction of adverse outcomes in older surgical patients? A population-based study. Age Ageing. 2021;50(3):802–808. doi:10.1093/ageing/afaa214.

25. Nghiem S, Afoakwah C, Scuffham P, Byrnes J. Hospital frailty risk score and adverse health outcomes: evidence from longitudinal record linkage cardiac data. Age Ageing. May 2021. doi:10.1093/ageing/afab073.

26. Nord M, Lyth J, Alwin J, Marcusson J. Costs and effects of comprehensive geriatric assessment in primary care for older adults with high risk for hospitalisation. BMC Geriatr. 2021;21(1):263. doi:10.1186/s12877-021-02166-1.

27. Charlson ME, Pompei P, Ales KL, MacKenzie CR. A new method of classifying prognostic comorbidity in longitudinal studies: Development and validation. J Chronic Dis. 1987;40(5):373–383. doi:10.1016/0021-9681(87)90171-8.

28. Rockwood K, Andrew M, Mitnitski A. A comparison of two approaches to measuring frailty in elderly people. Journals Gerontol Ser A. 2007;62(7):738–743. doi:10.1093/gerona/62.7.738.

29. Aliberti MJR, Szlejf C, Avelino-Silva VI, et al. COVID-19 is not over and age is not enough: Using frailty for prognostication in hospitalized patients. J Am Geriatr Soc. 2021;69(5):1116–1127. doi:10.1111/jgs.17146.

30. Ludvigsson JF, Appelros P, Askling J, et al. Adaptation of the Charlson comorbidity index for register-based research in Sweden. Clin Epidemiol. 2021;13:21–41. doi:10.2147/CLEP.S282475.

31. Kim DH. Measuring frailty in health care databases for clinical care and research. Ann Geriatr Med Res. 2020;24(2):62–74. doi:10.4235/agmr.20.0002.

32. Brundle C, Heaven A, Brown L, et al. Convergent validity of the electronic frailty index. Age Ageing. 2019;48(1):152–156. doi:10.1093/ageing/afy162.

33. Hollinghurst J, Housley G, Watkins A, Clegg A, Gilbert T, Conroy SP. A comparison of two national frailty scoring systems. Age Ageing. November 2020. doi:10.1093/ageing/afaa252.

34. Howlett SE, Rutenberg AD, Rockwood K. The degree of frailty as a translational measure of health in aging. Nat Aging. 2021;1(8):651–665. doi:10.1038/s43587-021-00099-3.

35. Singh I, Gallacher J, Davis K, Johansen A, Eeles E, Hubbard RE. Predictors of adverse outcomes on an acute geriatric rehabilitation ward. Age Ageing. 2012;41(2):242–246. doi:10.1093/ageing/afr179.

36. Gordon EH, Peel NM, Hubbard RE. The male-female health-survival paradox in hospitalised older adults. Maturitas. 2018;107:13–18. doi:10.1016/j.maturitas.2017.09.011.

37. Howlett SE, Rockwood MRH, Mitnitski A, Rockwood K. Standard laboratory tests to identify older adults at increased risk of death. BMC Med. 2014;12(1):171. doi:10.1186/s12916-014-0171-9.

38. Blodgett JM, Theou O, Howlett SE, Wu FCW, Rockwood K. A frailty index based on laboratory deficits in community-dwelling men predicted their risk of adverse health outcomes. Age Ageing. 2016;45(4):463–468. doi:10.1093/ageing/afw054.

39. Hägg S, Jylhävä J. Sex differences in biological aging with a focus on human studies. Suh Y, Tyler JK, eds. Elife. 2021;10:e63425. doi:10.7554/eLife.63425.

40. Hao Q, Zhou L, Dong B, Yang M, Dong B, Weil Y. The role of frailty in predicting mortality and readmission in older adults in acute care wards: a prospective study. Sci Rep. 2019;9(1):1207. doi:10.1038/s41598-018-38072-7.

41. Theou O, O’Connell MDL, King-Kallimanis BL, O’Halloran AM, Rockwood K, Kenny RA. Measuring frailty using self-report and test-based health measures. Age Ageing. 2015;44(3):471–477. doi:10.1093/ageing/afv010.

42. Hubbard RE, Peel NM, Samanta M, Gray LC, Mitnitski A, Rockwood K. Frailty status at admission to hospital predicts multiple adverse outcomes. Age Ageing. 2017;46(5):801–806. doi:10.1093/ageing/afx081.

43. Wou F, Gladman JRF, Bradshaw L, Franklin M, Edmans J, Conroy SP. The predictive properties of frailty-rating scales in the acute medical unit. Age Ageing. 2013;42(6):776–781. doi:10.1093/ageing/aft055.

44. Kansagara D, Englander H, Salanitro A, et al. Risk prediction models for hospital readmission: a systematic review. JAMA. 2011;306(15):1688–1698. doi:10.1001/jama.2011.1515.

45. Navathe AS, Zhong F, Lei VJ, et al. Hospital readmission and social risk factors identified from physician notes. Health Serv Res. 2018;53(2):1110–1136. doi:10.1111/1475-6773.12670.

46. Mudge AM, Hubbard RE. Frailty: mind the gap. Age Ageing. 2018;47(4):508–511. doi:10.1093/ageing/afx193.

47. Grundberg Å, Ebbeskog B, Abrandt Dahlgren M, Religa D. How community-dwelling seniors with multimorbidity conceive the concept of mental health and factors that may influence it: a phenomenographic study. Int J Qual Stud Health Well-being. 2012;7(1):19716. doi:10.3402/qhw.v7i0.19716.

