## Supplementary data for "Development of an electronic frailty index for hospitalized geriatric patients in Sweden"

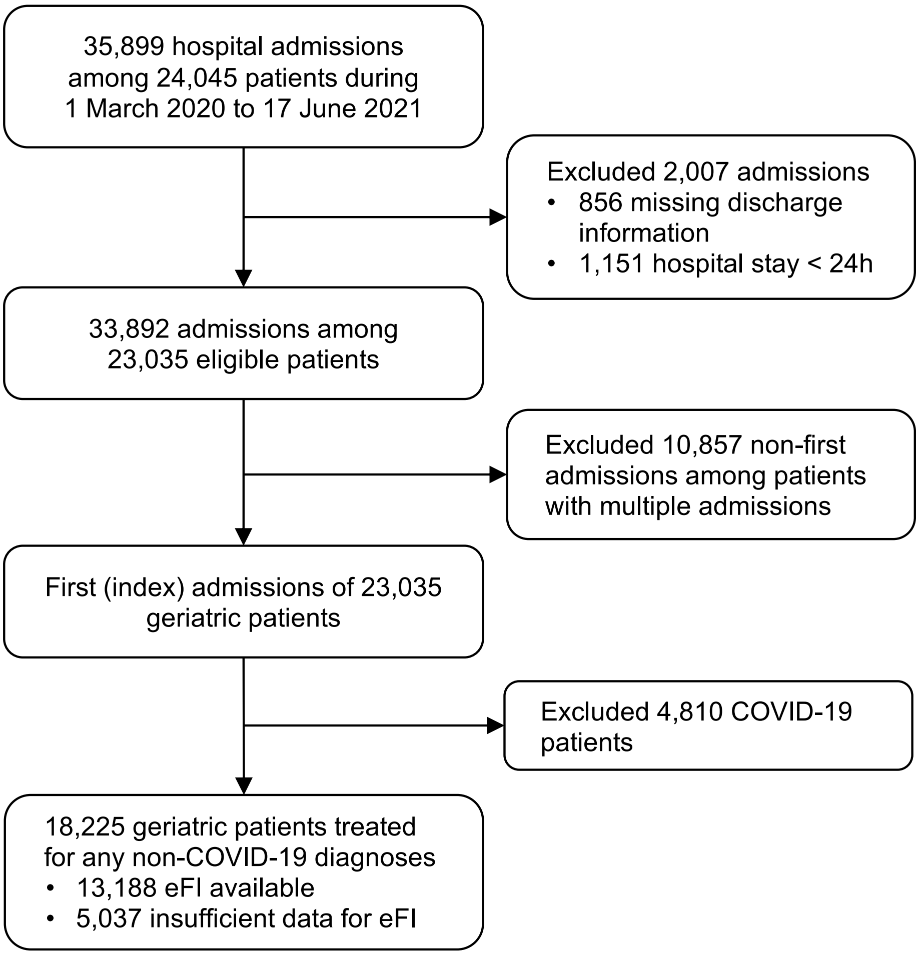

**Supplementary Figure 1.** Flowchart of sample selection

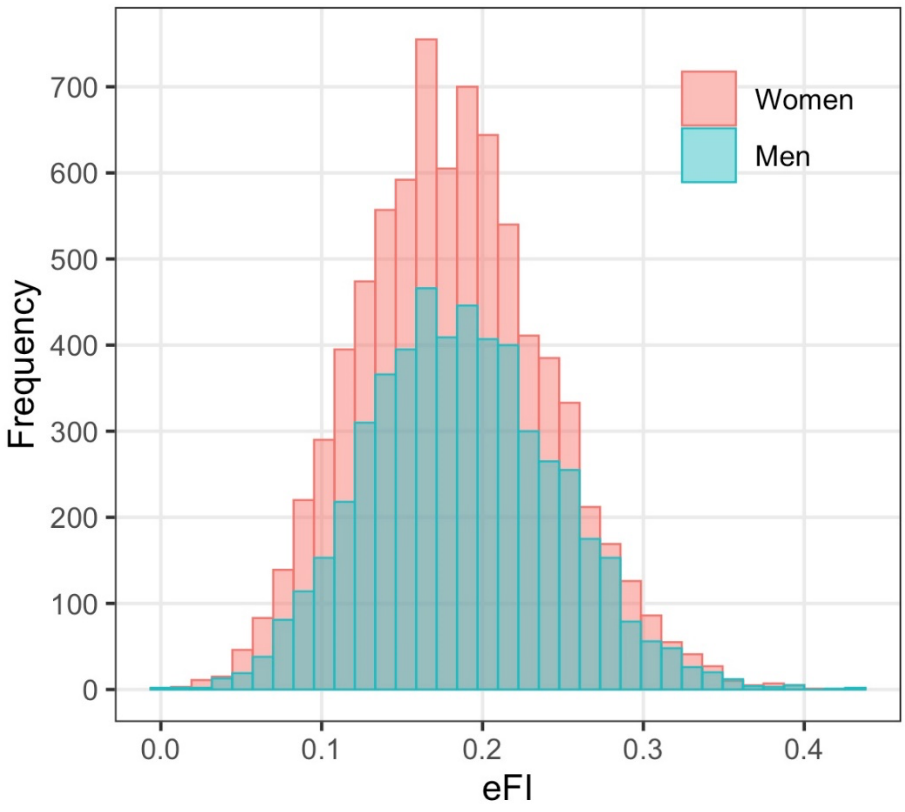

**Supplementary Figure 2.** Distribution of the electronic frailty index by sex (n = 13,188)

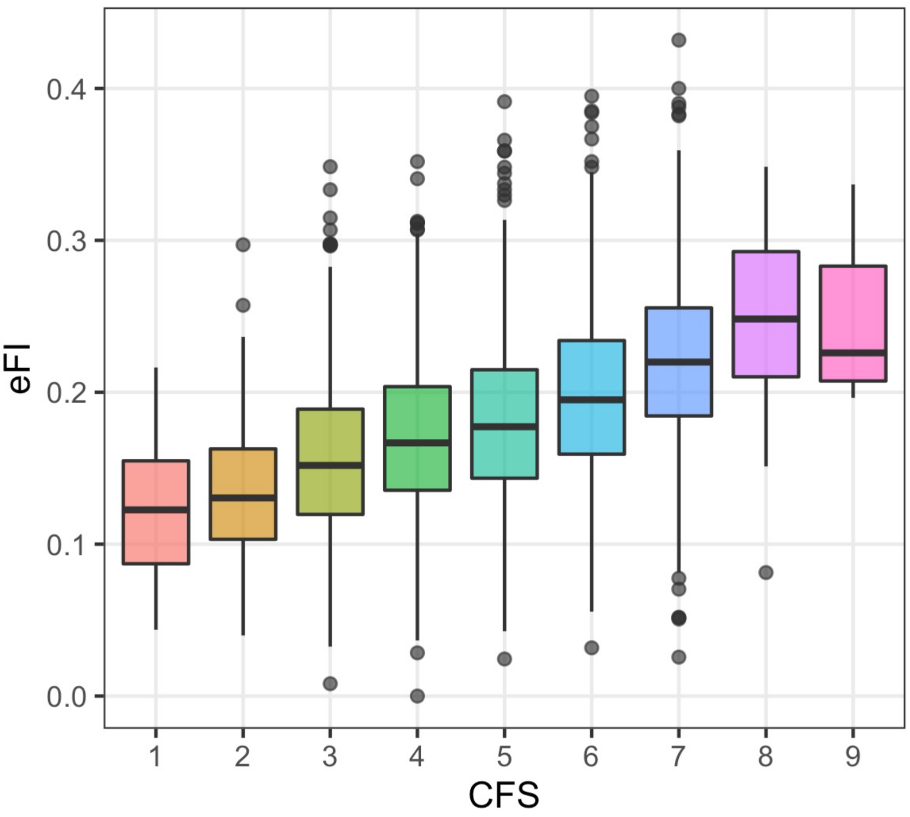

**Supplementary Figure 3.** Box plot of the electronic frailty index by levels of the Clinical Frailty Scale (n = 4,945)

**Supplementary Table 1.** List of the 48 deficit items and coding for construction of the electronic frailty index

| No. | Deficits | Coding |
|---|---|---|
| ***Diagnosis code-based deficits*** | | |
| 1 | Anemia | 1 = Any of the ICD-10 codes: D50.0, D50.8, D50.9, D51–D53 |
| 2 | Asthma | 1 = Any of the ICD-10 codes: J45 |
| 3 | Atrial fibrillation | 1 = Any of the ICD-10 codes: I48 |
| 4 | Cancer | 1 = Any of the ICD-10 codes: C00–C97 |
| 5 | Chronic pain | 1 = Any of the ICD-10 codes: F45.4, M08.1, M25.5, M43.2–M43.6, M45, M46.1, M46.3, M46.4, M46.9, M47, M48.0, M48.1, M48.8, M48.9, M50.8, M50.9, M51, M53.1–M53.3, M53.8, M53.9, M54, M60.8, M60.9, M63.3, M79.0–M79.2, M79.6, M79.7, M96.1 |
| 6 | Congestive heart failure | 1 = Any of the ICD-10 codes: I09.9, I11.0, I13.0, I13.2, I25.5, I42.0, I42.5–I42.9, I43, I50, P29.0 |
| 7 | Coronary atherosclerosis and other heart disease | 1 = Any of the ICD-10 codes: I20, I24, I25, Z95.1 |
| 8* | Dementia | 1 = Any of the ICD-10 codes: F00-F03, F05.1, G30, G31.1 |
| 9 | Depression | 1 = Any of the ICD-10 codes: F20.4, F31.3–F31.5, F32, F33, F34.1, F41.2, F43.2 |
| 10 | Diabetes | 1 = Any of the ICD-10 codes: E10–E14 |
| 11 | Dizziness or vertigo | 1 = Any of the ICD-10 codes: H81.0, H81.1, H81.2, H81.3, H81.4, H81.8, H81.9, H82.9, H83.0, H83.1, H83.2, H83.8, H83.9, R42 |
| 12 | Dyspnea | 1 = Any of the ICD-10 codes: R06.0, R06.2–R06.4, R06.8, R06.9 |
| 13 | Fragility fracture | 1 = Any of the ICD-10 codes: M48.5, M80.0, M80.8, M84.3–M84.7, S02, S12, S22, S32, S42, S52, S62, S72, S82, S92 |
| 14* | Hypertension | 1 = Any of the ICD-10 codes: I10–I13, I15 |
| 15 | Hypotension/syncope | 1 = Any of the ICD-10 codes: I95, R55 |
| 16 | Liver disease | 1 = Any of the ICD-10 codes: B18, I85, I86.4, I98.2, K70, K71.1, K71.3–K71.5, K71.7, K72–K74, K76.0, K76.2–K76.9, Z94.4 |
| 17 | Myocardial infarction | 1 = Any of the ICD-10 codes: I21, I22, I25.2 |
| 18 | Osteoporosis | 1 = Any of the ICD-10 codes: M80, M81 |
| 19 | Parkinsonism and tremor | 1 = Any of the ICD-10 codes: G20, G21, G22, G25 |
| 20 | Peptic ulcer | 1 = Any of the ICD-10 codes: K25–K28 |
| 21 | Peripheral vascular disease | 1 = Any of the ICD-10 codes: I70, I71, I73.1, I73.8, I73.9, I77.1, I79.0, I79.2, K55.1, K55.8, K55.9, Z95.8, Z95.9 |
| 22 | Pulmonary disease | 1 =Any of the ICD-10 codes: I27.8, I27.9, J40–J47, J60–J67, J68.4, J70.1, J70.3 |
| 23 | Renal disease | 1 = Any of the ICD-10 codes: I12.0, I13.1, N03.2–N03.7, N05.2–N05.7, N18, N19, N25.0, Z49.0–Z49.2, Z94.0, Z99.2 |
| 24 | Rheumatoid arthritis or osteoarthritis | 1 = Any of the ICD-10 codes: M05, M06, M15–M19, M31.5, M32-M34, M35.1, M35.3, M36.0 |
| 25 | Skin ulcer | 1 = Any of the ICD-10 codes: L89, L97, L98.4 |
| 26 | Stroke or transient ischemic attack | 1 = Any of the ICD-10 codes: G45.0–G45.3, G45.8–G45.9, H34.1, I60, I61, I63, I64 |
| 27 | Thyroid disease | 1 = Any of the ICD-10 codes: E00–E07, E89.0, R94.6 |
| 28 | Urinary system disease | 1 = Any of the ICD-10 codes: F52.9, N13, N20, N21, N23, N25, N27, N28, N31, N32, N35, N36, N39, R30, R31, R33, R33.9, R34, R35, R36.0, R36.9, R39, R80, R82.0–R82.3, R82.5, R93.4, R94.4, Z43.5, Z43.6, Z46.6, Z87.4, Z93.5, Z93.6, Z96.0 |
| 29 | Valvular disease | 1 = Any of the ICD-10 codes: A52.0, I05–I08, I09.1, I09.8, I34–I39, Q23.0–Q23.3, Z95.2–Z95.4 |
| ***Functioning and other health indicators*** | | |
| 30 | Activity limitation | Norton: 0 = Ambulant; 1/3 = Walks with help; 2/3 = Chairbound; 1 = Bedfast<br>MNA: 0 = Goes out; 1/3 = Able to get out of bed/chair but does not go out; 1 = Bed or chair bound<br>(Take the maximum of either one) |
| 31 | Cognitive impairment | Downton: 0 = Oriented; 1 = Confused<br>Norton: 0 = Alert or Apathetic; 1 = Confused or Stuporous<br>(Take the maximum of either one) |
| 32 | Falls | Downton: 0 = No; 1 = Yes |
| 33 | Food intake status | MNA: 0 = No decrease in food intake; 0.5 = Moderate decrease in food intake; 1 = Severe decrease in food intake<br>Norton: 0 = Normal portion; 1/3 = 3/4 portion; 2/3 = 1/2 portion; 1 = 1/2 portion<br>(Take the maximum of either one) |
| 34 | General condition | Norton: 0 = Good; 1/3 = Fair; 2/3 = Poor; 1 = Very Bad |
| 35 | Incontinence | Norton: 0 = None; 1/3 = Occasional; 2/3 = Urinary or fecal; 1 = Urinary and fecal<br>Barthel: 0 = Continent in both bowel and bladder; 1/3 = Occasional accident in bowel or bladder; 2/3 = Incontinent in bowel or bladder; 1 = Incontinent in both bowel and bladder<br>(Take the maximum of either one) |
| 36 | Mobility | Norton: 0 = No; 1/3 = Slightly Impaired; 2/3 = Very Limited; 1 = Immobile |
| 8* | Neuropsychological problems | MNA: 0 = No neuropsychological problems; 1 = Mild/ severe dementia or depression |
| 37 | Oral health | ROAG: 0 = none of the nine items is 2 or above; 0.5 = any of the nine items equals 2; 1 = any of the nine items equals 3 |
| 38 | Sensory impairment | Downton: 0 = No sensory impairment; 1/3 = One impairment; 2/3 = Two impairments; 1 = Three impairments |
| 39 | Weight loss | MNA: 0 = No weight loss/don't know; 0.5 = 1-3 kg during last 3 months; 1 = >3 kg during last 3 months |

**Supplementary Table 1 (continued).**

| No. | Deficits | Coding |
|---|---|---|
| ***Laboratory/anthropometric measures*** | | |
| 40 | C-reactive protein | 0 = <3 mg/L; 0.5 = 3–10 mg/L; 1 = >10 mg/L |
| 41 | Creatinine | 0 = 60–110 µmol/L for men and 45–90 µmol/L for women; 1 = <60 µmol/L or >110 µmol/L for men, and <45 µmol/L or >90 µmol/L for women |
| 42 | Glucose | 0 = 4.4–11.1 mmol/L; 1 = <4.4 or >11.1 mmol/L |
| 43 | Hemoglobin | 0 = 135–180 g/L for men and 120–160 g/L for women; 1 = <135 or >180 g/L for men, and <120 or >160 g/L for women |
| 14* | High systolic/diastolic blood pressure | 0 = systolic<140 or diastolic<90; 0.5 = 140≤systolic<180 or 90≤diastolic<100; 1 = systolic≥180 or diastolic≥100 |
| 44 | Obesity | 0 = BMI<30; 1 = BMI≥30 |
| 45 | Potassium | 0 = 3.5–6 mmol/L; 1 = <3.5 or >6 mmol/L |
| 46 | Pulse | 0 = 60–99 bpm; 1 = <60 or >99 bpm |
| 47 | Sodium | 0 = 136–142 mmol/L; 1 = <136 or >142 mmol/L |
| 48 | Underweight | 0 = BMI ≥18.5; 1 = BMI <18.5 |

*Note:* The eFI was calculated only if a patient had (i) ≥30 non-missing deficit items and (ii) at least half of the functional and/or laboratory items being non-missing. Norton = Norton scale for pressure ulcer risk assessment; Downton = Downton fall risk assessment scale; MNA = Mini-Nutritional Assessment Scale; Barthel = Barthel scale for activities of daily living; ROAG = Revised Oral Assessment Guide
* Overall deficit score=maximum of diagnosis code-based deficit and functioning/ lab deficits

**Supplementary Table 2.** Sample characteristics by availability of the electronic frailty index

| Characteristic | All patients (n = 18,225) | eFI available (n = 13,188) | Insufficient data for eFI[a] (n = 5,037) | $p^b$ |
|---|---|---|---|---|
| Age, year, mean ± SD | 83.1 ± 8.4 | 83.1 ± 8.4 | 82.9 ± 8.3 | 0.19 |
| Age category, n (%) | | | | 0.39 |
| <65 years | 287 (1.6) | 215 (1.6) | 72 (1.4) | |
| 65–74 years | 2,665 (14.6) | 1,917 (14.5) | 748 (14.9) | |
| 75–84 years | 6,827 (37.5) | 4,909 (37.2) | 1,918 (38.1) | |
| 85–94 years | 7,037 (38.6) | 5,103 (38.7) | 1,934 (38.4) | |
| ≥95 years | 1,409 (7.7) | 1,044 (7.9) | 365 (7.2) | |
| Men, n (%) | 7,309 (40.1) | 5,245 (39.8) | 2,064 (41.0) | 0.14 |
| CFS score, median [IQR] | 5 [4, 6] | 5 [4, 6] | 5 [4, 6] | 0.73 |
| CFS category, n (%) | | | | 0.38 |
| 1–3 | 957 (16.6) | 830 (16.8) | 127 (15.8) | |
| 4–5 | 2,196 (38.2) | 1,869 (37.8) | 327 (40.6) | |
| 6–9 | 2,598 (45.2) | 2,246 (45.4) | 352 (43.7) | |
| Missing | 12,474 | 8,243 | 4,231 | |
| HFRS, median [IQR] | 2.8 [1.4, 5.0] | 2.8 [1.4, 5.0] | 2.8 [1.4, 5.0] | 0.91 |
| HFRS category, n (%) | | | | 0.98 |
| Low risk (<5) | 13,552 (74.4) | 9,811 (74.4) | 3,741 (74.3) | |
| Intermediate risk (5–15) | 4,589 (25.2) | 3,316 (25.1) | 1,273 (25.3) | |
| High risk (>15) | 84 (0.5) | 61 (0.5) | 23 (0.5) | |
| CCI, median [IQR] | 1 [0, 2] | 1 [0, 2] | 1 [0, 2] | <0.001 |
| In-hospital mortality, n (%) | 266 (1.5) | 183 (1.4) | 83 (1.6) | 0.22 |
| Discharged to home, n (%) | 14,407 (79.1) | 10,448 (79.2) | 3,959 (78.6) | 0.38 |
| 30-day readmission[c], n (%) | 1,521 (10.6) | 1,114 (10.7) | 407 (10.3) | 0.53 |
| Length of stay, day, median [IQR] | 6.6 [4.6, 8.8] | 6.7 [4.7, 8.9] | 6.4 [4.4, 8.7] | <0.001 |

*Note*: CCI, Charlson Comorbidity Index; CFS, Clinical Frailty Scale; eFI, electronic frailty index; HFRS, Hospital Frailty Risk Score; IQR, interquartile range; SD, standard deviation

[a] Patients with insufficient data for eFI had <30 non-missing deficit items, and/or had <9 (out of 18) non-missing functional and laboratory items

[b] *P*-values for comparison between patients with and without sufficient data for eFI, based on *t*-tests or Mann-Whitney *U* tests for continuous variables, and $\chi^2$ tests for categorical variables

[c] Only patients discharged to home after the first admission were considered for 30-day readmission

**Supplementary Table 3.** Spearman's correlations between frailty and comorbidity measures (n = 4,945)

| *ρ* | CFS | HFRS | CCI | eFI |
|---|---|---|---|---|
| CFS | 1 | | | |
| HFRS | 0.244 | 1 | | |
| CCI | 0.096 | 0.140 | 1 | |
| eFI | 0.420 | 0.289 | 0.368 | 1 |

*Note*: CFS, Clinical Frailty Scale; HFRS, Hospital Frailty Risk Score; CCI, Charlson Comorbidity Index; eFI, electronic frailty index

**Supplementary Table 4.** Logistic regression models for the associations between the electronic frailty index and in-hospital mortality stratified by the admitting clinics

| Admitting clinics | In-hospital deaths / No. of patients (%) | Adjusted OR per 10% increase in eFI (95% CI)[a] | AUC |
|---|---|---|---|
| Clinic 1 | 30/1,916 (1.6) | 7.42 (3.79, 15.0)* | 0.861 |
| Clinic 2 | 8/1,052 (0.8) | 6.37 (2.03, 20.6)* | 0.837 |
| Clinic 3 | 25/1,978 (1.3) | 3.53 (1.80, 6.93)* | 0.792 |
| Clinic 4 | 16/1,137 (1.4) | 4.87 (2.08, 11.8)* | 0.745 |
| Clinic 5 | 8/999 (0.8) | 3.39 (1.04, 10.8)* | 0.827 |
| Clinic 6 | 13/1,904 (0.7) | 7.02 (2.81, 18.1)* | 0.871 |
| Clinic 7 | 1/234 (0.4) | Not estimable | - |
| Clinic 8 | 26/2,542 (1.0) | 4.41 (2.37, 8.28)* | 0.848 |
| Clinic 9 | 56/1,426 (3.9) | 5.31 (3.41, 8.41)* | 0.808 |

*Note*: AUC, area under the receiver operating characteristic curve; CI, confidence interval; eFI, electronic frailty index; OR, odds ratio

[a] All the listed models were multivariate logistic regression models adjusted for age and sex.

* $p<0.05$

**Supplementary Table 5.** Logistic regression models for the associations between frailty and comorbidity measures and 30-day readmission (n = 10,448)[a]

| Model | 30-day readmission | |
|---|---|---|
| | Adjusted OR (95% CI) | AUC |
| **eFI** | | |
| Continuous (per 10% increase) | 1.50 (1.35, 1.68)* | 0.592 |
| Categorical | | |
| Fit (≤0.15) | 1 (Ref.) | 0.591 |
| Mild frailty (>0.15–0.2) | 1.29 (1.10, 1.53)* | |
| Moderate frailty (>0.2–0.25) | 1.85 (1.56, 2.20)* | |
| Severe frailty (>0.25) | 1.82 (1.46, 2.25)* | |
| **CFS (n = 4,945)**[b] | | |
| Continuous (per point increase) | 1.14 (1.06, 1.23)* | 0.586 |
| Categorical | | |
| 1–3 | 1 (Ref.) | 0.585 |
| 4–5 | 1.20 (0.88, 1.67) | |
| 6–9 | 1.58 (1.17, 2.17)* | |
| **HFRS** | | |
| Continuous (per point increase) | 1.03 (1.01, 1.05)* | 0.564 |
| Categorical | | |
| Low risk (<5) | 1 (Ref.) | 0.561 |
| Intermediate risk (5–15) | 1.16 (1.01, 1.34)* | |
| High risk (>15) | 0.79 (0.19, 2.23) | |
| **CCI** | | |
| Continuous (per point increase) | 1.12 (1.08, 1.17)* | 0.578 |

*Note*: AUC, area under the receiver operating characteristic curve; CCI, Charlson Comorbidity Index; CFS, Clinical Frailty Scale; CI, confidence interval; eFI, electronic frailty index; HFRS, Hospital Frailty Risk Score; OR, odds ratio

[a] All the listed models were multivariate logistic regression models adjusted for age and sex. Only patients discharged to home after the first admission were included in the 30-day readmission analysis. eFI, CFS, and HFRS were used as both continuous and categorical variables in separate models, while CCI was used as continuous variable only.

[b] Sample size was smaller in analysis of CFS due to missing data

* $p<0.05$

**Supplementary Table 6.** Linear regression models for the associations between frailty and comorbidity measures and length of stay (n = 13,188)[a]

| Model | Length of stay | |
|---|---|---|
| | Adjusted *β* (95% CI) | $R^2$ |
| **eFI** | | |
| Continuous (per 10% increase) | 2.00 (1.88, 2.13)* | 7.21% |
| Categorical | | |
| Fit (≤0.15) | 0 (Ref.) | 6.05% |
| Mild frailty (>0.15–0.2) | 1.18 (1.00, 1.36)* | |
| Moderate frailty (>0.2–0.25) | 2.13 (1.93, 2.33)* | |
| Severe frailty (>0.25) | 3.22 (2.98, 3.47)* | |
| **CFS (n = 4,945)**[b] | | |
| Continuous (per point increase) | 0.54 (0.46, 0.62)* | 3.50% |
| Categorical | | |
| 1–3 | 0 (Ref.) | 3.06% |
| 4–5 | 0.93 (0.57, 1.29)* | |
| 6–9 | 2.03 (1.68, 2.38)* | |
| **HFRS** | | |
| Continuous (per point increase) | 0.21 (0.19, 0.23)* | 2.39% |
| Categorical | | |
| Low risk (<5) | 0 (Ref.) | 1.61% |
| Intermediate risk (5–15) | 1.21 (1.04, 1.38)* | |
| High risk (>15) | 1.95 (0.86, 3.04)* | |
| **CCI** | | |
| Continuous (per point increase) | 0.21 (0.16, 0.25)* | 0.64% |

*Note*: CCI, Charlson Comorbidity Index; CFS, Clinical Frailty Scale; CI, confidence interval; eFI, electronic frailty index; HFRS, Hospital Frailty Risk Score

[a] All the listed models were multivariate linear regression models adjusted for age and sex. eFI, CFS, and HFRS were used as both continuous and categorical variables in separate models, while CCI was used as continuous variable only.

[b] Sample size was smaller in analysis of CFS due to missing data

* $p<0.05$
